# The characteristics of HIV-positive patients with mild/asymptomatic and moderate/severe course of COVID-19 disease – a report from Central and Eastern Europe

**DOI:** 10.1101/2020.10.28.20221226

**Authors:** Justyna D. Kowalska, Kerstin Kase, Anna Vassilenko, Arjan Harxhi, Botond Lakatos, Gordana Dragović Lukić, Antonija Verhaz, Nina Yancheva, Florentina Dumitrescu, David Jilich, Ladislav Machala, Agata Skrzat-Klapaczyńska, Raimonda Matulionyte

## Abstract

**Background:** There is currently no evidence suggesting that COVID-19 takes a different course in HIV-positive patients on antiretroviral treatment compared to the general population. However, little is known about the relation between specific HIV-related factors and the severity of the COVID-19 disease.

**Methods:** We performed a retrospective analysis of cases collected through an on-line survey distributed by the Euroguidelines in Central and Eastern Europe Network Group. In statistical analyses characteristics of HIV-positive patients asymptomatic/moderate and moderate/severe course were compared.

**Results:** In total 34 HIV-positive patients diagnosed with COVID-19 were reported by 12 countries (Estonia, Czech Republic, Lithuania, Albania, Belarus, Romania, Serbia, Bosnia and Herzegovina, Poland, Russia, Hungary, Bulgaria). Asymptomatic courses of COVID-19 were reported in four (12%) cases, 11 (32%) patients presented with mild disease not requiring hospitalization, moderate disease with respiratory and/or systemic symptoms was observed in 14 (41%) cases, and severe disease with respiratory failure was found in five (15%) patients. The only HIV-related characteristics differentiating a moderate/severe course of the disease from asymptomatic/mild disease course was the use of or PI or NNRTI as part of the cART regimen (40.0% vs. 5.3%, p=0.0129 for PI and 31.6 % vs. 0.0%, p= 0.0239 for NNRTI).

**Conclusions:** In our analyses HIV viral suppression and immunological status were not playing a role in the course of COVID-19 disease. On the contrary the cART regimen could contribute to severity of SARS-CoV-2 infection. Large and prospective studies are necessary to further investigate this relations.

## Introduction

The ongoing COVID-19 pandemic may affect many aspects of HIV care, from individual health to the continuation of antiretroviral treatment and its sustainability (1). It is currently assumed that there is no increased risk of infection, or of a more severe course of the COVID-19 disease among HIV-positive patients on combined antiretroviral therapy (cART) (2). This condition may not be well fulfilled in many Central and Eastern Europe (CEE) countries, where the cascade of care is insufficient and there are many barriers to accessing cART (3).

It is also interpreted that the same risk factors play a role in HIV-positive persons as they do in the general population, namely older age, immunosuppression, and certain comorbidities (4, 5). However, as the HIV-positive population is ageing a higher prevalence of comorbidities compared to the general population is observed (6). Extrapolating from these observations we could expect the HIV-positive population to be disproportionately affected by COVID-19. On the contrary first observations of the confirmed SARS-CoV-2 cases did not present any significant difference in the course of COVID-19 for HIV positive persons (7-11). Some recent publications seems to present increase risk of death among HIV-positive persons, but available data does not allow for final assessment of associations with current immune status, viral suppression or antiretroviral therapy (12, 13). Each of these is important for investigation of the HIV and SARS-CoV-2 interplay in death causality. The relation between the severity of the COVID-19 disease and HIV specific factors, such as particular cART components, was possible only in few studies (14). In our study, we analyze the risk factors for moderate/severe courses of COVID-19 among HIV-positive patients reported by CEE countries.

## Material and methods

The Euroguidelines in Central and Eastern Europe Countries (ECEE) Network Group was established in February 2016 to review the standards of care for HIV infection and viral hepatitis in the region. The Network consists of key experts from the field who are actively involved in HIV care. In March 2020, the group decided to collect and analyze all known cases of COVID-19 in HIV-positive patients in the CEE. Data were collected through an anonymized on-line questionnaire. The information collected included demographics, time since HIV diagnosis, baseline and most recent HIV viral load, nadir and most recent CD 4+ cell count, cART status and components, and comorbidities and coinfections. Clinical courses of COVID-19 were defined as asymptomatic (no symptoms throughout the course of infection), mild disease (symptomatic patients not requiring hospitalization), moderate disease (stable symptomatic patients with respiratory and/or systemic symptoms requiring hospitalization), and severe disease (clinically unstable patients with respiratory failure requiring hospitalization). For the purpose of the analysis, the patients were categorized into asymptomatic/mild and moderate/severe COVID-19 disease. Non-parametric tests were used in the statistical analyses for groups comparison as appropriate.

## Results

In total, 34 HIV-positive patients diagnosed with COVID-19 between March 11 and June 26, 2020 were reported by 12 countries (Estonia, Czech Republic, Lithuania, Albania, Belarus, Romania, Serbia, Bosnia and Herzegovina, Poland, Russia, Hungary, Bulgaria). In general 29.4 % of patients were women, median age was 40.5 years, median body mass index 26.0 kg/m3 and 19 (55.9 %) patients had comorbidities. Most of the patients acquired HIV through sexual contacts (79.4 %) and median time since HIV diagnosis was 5 years. Most of the patients were on cART (82.3 %) but only half of them (54.5%) had HIV RNA undetectable. Most recent median CD4+ count was 557 cells/mm^3^.

Asymptomatic courses of COVID-19 were reported in four (12 %) cases, 11 (32 %) patients presented with mild disease not requiring hospitalization Moderate disease with respiratory and/or systemic symptoms was observed in 14 (41 %) cases, and severe disease with respiratory failure in five (15 %) patients.

The HIV-related characteristics of patients with asymptomatic/mild course of COVID-19 were comparable to those with moderate/severe course of COVID-19, except for the use of protease inhibitors (PIs) and non-nucleoside reverse transcriptase inhibitors (NNRTIs) in cART regimen. Another notable difference was for medical interventions, such as intensive care unit stay, need for mechanical ventilation and using COVID-19 specific treatment, as expected it was higher in moderate/severe course of COVID-19 group, Table 1. In terms of COVID-19 treatment six patients received hydroxychloroquine, two lopinavir/ritonavir, one azithromycin, one favipravir with tocilizumab in moderate/severe group. Only one patient received COVID-19 treatment in asymptomatic/mild group (chloroquine).

**Table 1.**
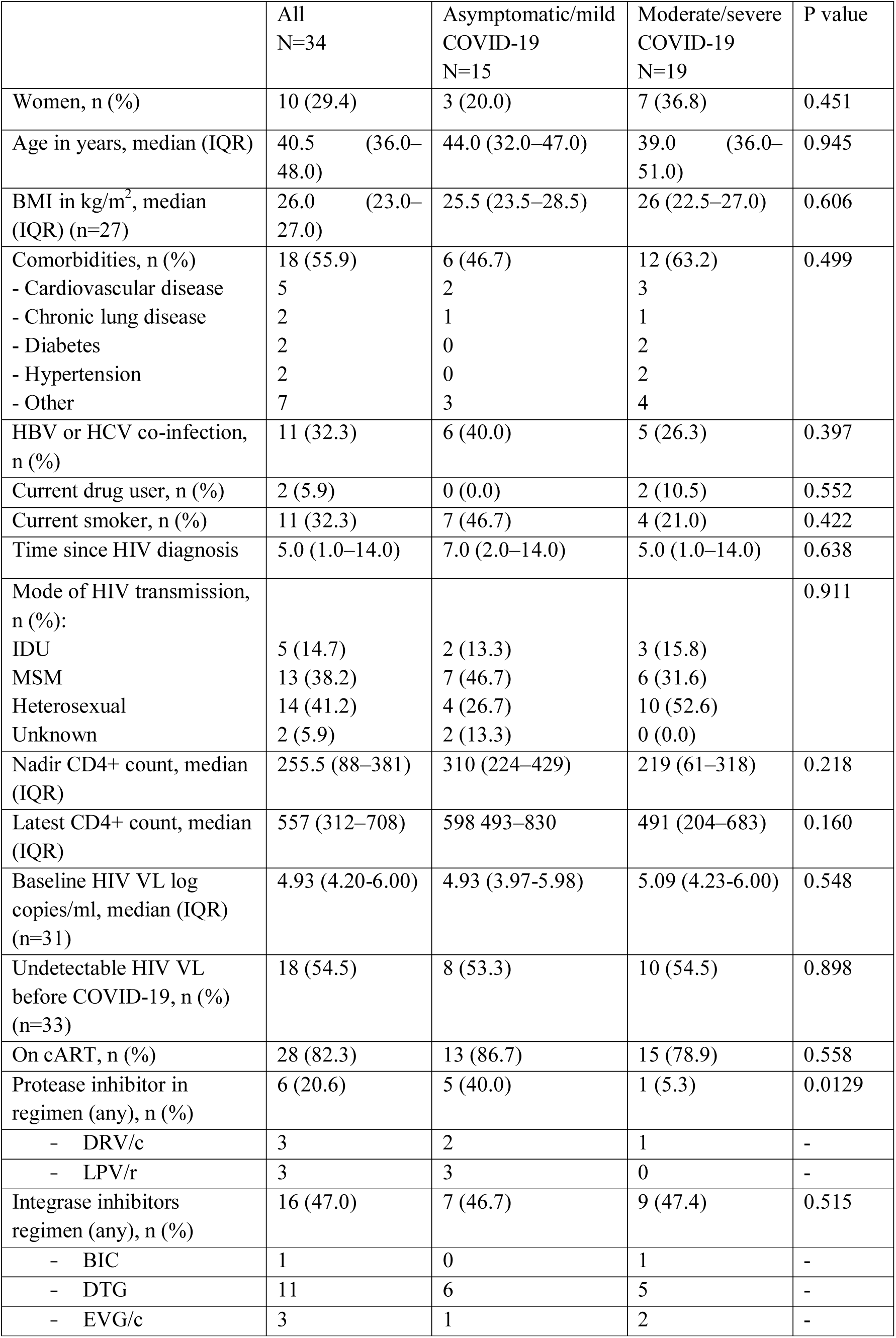
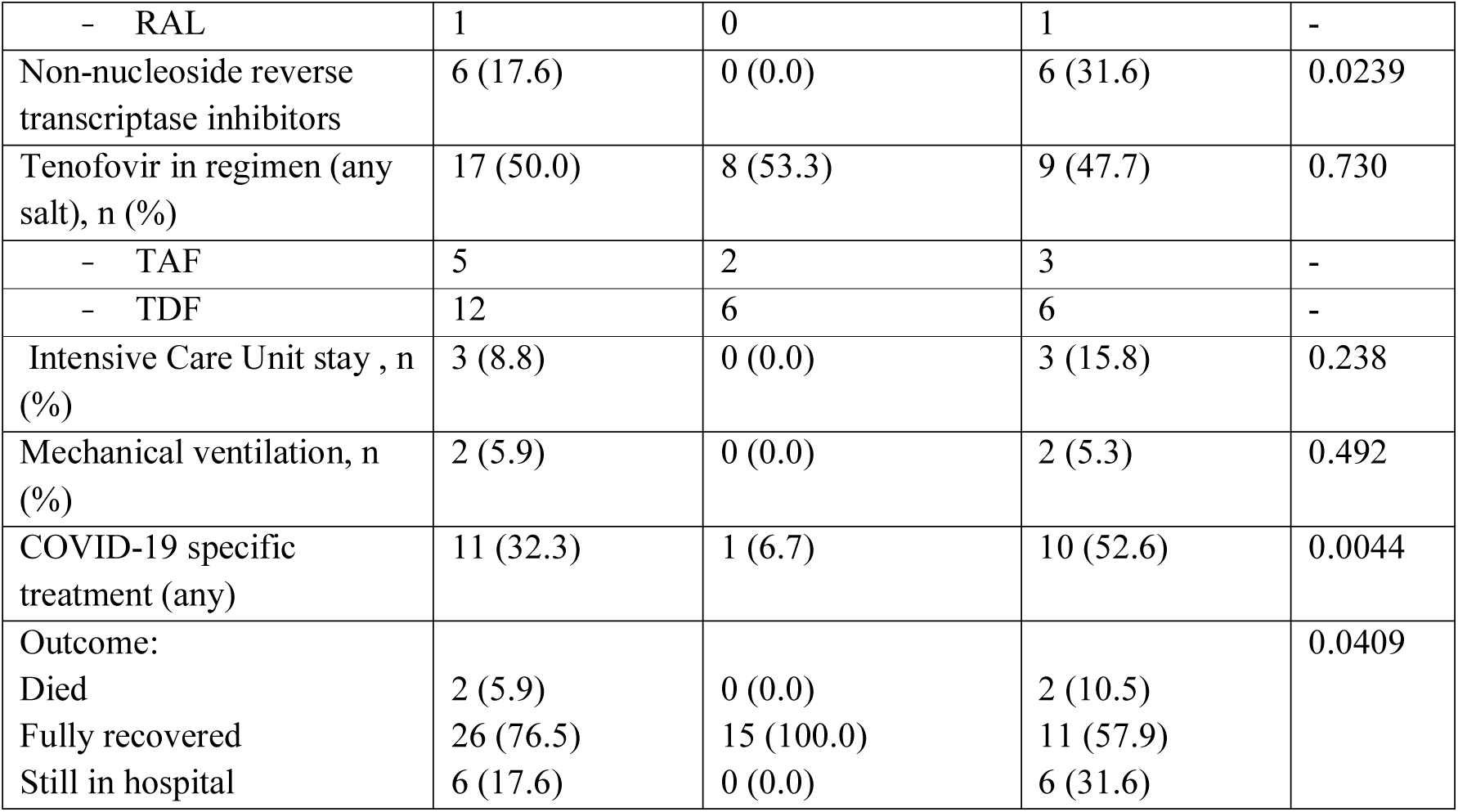
Baseline characteristics of HIV-positive patients with COVID-19 diagnosis.

## Discussion

Our analyses of 34 case series did not present any notable difference between HIV-positive patients with asymptomatic/mild and moderate/severe COVID-19 disease course. indicated that patients with well controlled HIV infection does not present an increased risk of poorer COVID-19 prognosis, but this cannot be extrapolated on uncontrolled HIV infection (12). This indicated the importance on reporting on the HIV infection and COVID-19 interplay in patients not on cART. To the best our knowledge this is the first report on COVID-19 among HIV-positive persons in CEE countries, where the cascade of HIV care remains an important public health problem, with lower rates of HIV-positive patients receiving cART (1, 3). In fact 82.6 % of patients in our study were on cART and only 54.5 % of these had undetectable viral load prior to COVID-19 diagnosis. Although we did not find undetectable viral load to have a protective effect against severe COVID-19 in this group of patients. Several of the studies published so far have not shown increased risk of severe COVID-19 outcomes by HIV status, or in relation to specific HIV-related characteristics (7-10, 15). However few of these studies were investigating factors potentially associated with moderate/severe courses of COVID-19 within the HIV population. A recent systematic review that analyzed potential risk factors confirmed that older age, comorbidities – as in the uninfected HIV population – and no specific HIV-related factors, may contribute to more severe outcomes in a setting where 98 % of patients are on cART, highlighting the significance of ensuring cART supply and adherence (5). The ISARIC CCP-UK study suggested a 63 % increased risk of day-28 mortality for HIV-positive patients (n=115) as compared with HIV-negative patients (n=47 979) (13). However this study did not have details of the CD4 lymphocyte count, ART history and HIV viral load.

We have observed the difference suggesting that the cART regimen may play a role in the course of SARS-CoV-2 related disease, yet this finding needs to be interpreted with cautious due to many limitations of this work. A recent study by del Amo et al. of 236 HIV-positive persons with COVID-19 diagnosis and receiving cART in Spain, suggested a decreased risk of COVID-19 and hospitalization among persons on tenofovir disoproxil with emtricitabine as compared to other NRTI components (16). This study did not analyze other cART components. We did not see the same associations in our study, however due to a small sample we were unable to disentangle the effects of different forms of tenofovir (17). Certainly there is a risk of bias per indication reflecting that ‘healthier patients” would stay on more toxic regimen containing tenofovir disoproxil, while patients with comorbidities would be switched or prescribed tenofovir alafenamide. Such confounding should be taken into account in COVID-19 and HIV research (18). As for other cART regimens in our study the most common third component in the regimen was integrase inhibitor (47.0 %), followed by protease inhibitors (20.6 %) and non-nucleoside reverse transcriptase inhibitors (17.6 %). The difference between COVID-19 severity group was observed for the latter two.

It has been indicated by other studies that neither tenofovir nor darunavir protect HIV-positive persons from acquiring SARS-CoV-2 (8, 19). In parallel, no benefit was observed, beyond standard care, for lopinavir use in the treatment of SARS-CoV-2 infection (20). However modest antiviral effects may not translate into susceptibility to SARS-CoV-2 infection or its mortality, but could contribute to the course of the COVID-19 disease expressed by length of hospital stay or oxygen therapy demand.

As our work is based on case series, there are important limitations to be considered when making conclusions from the results. First, the low number of asymptomatic cases indicates that people without symptoms do not seek medical assistance and therefore remain underreported. We cannot exclude the possibility that the pattern of cART regimen prescription may differ in this population, and thus influence our final results. Only analyses of large and prospectively designed HIV cohorts could overcome this limitation.

In our analyses HIV viral suppression and immunological status, as well as any other HIV-related factor, were not playing a role in the course of COVID-19 disease. On the contrary the cART regimen could contribute to severity of SARS-CoV-2 infection. Large and prospective studies are necessary to further investigate this relations.

## Data Availability

Data available upon request.

## Declarations

Ethics approval was obtained.

### Consent for publication

all co-authors give their consent for this work to be published.

### Data and materials

available upon request.

### Authors’ contributions

all co-authors contributed equally to this work.

